# Prevalence and associated factors of suicidal ideation and suicide attempts in adolescents attending St. Paul’s Hospital Millennium Medical College, Pediatric Clinic in Addis Ababa, Ethiopia, 2023

**DOI:** 10.1101/2025.03.18.25324208

**Authors:** Dawit Sintayehu, Tigist Zerihun, Tigist Bacha

## Abstract

**Background:** Suicide among adolescents is a significant global public health concern. This study aimed to assess the prevalence and associated factors of suicidal ideation and attempts among adolescents attending care at St. Paul’s Hospital Millennium Medical College, Pediatrics Department, Addis Ababa, Ethiopia.

**Methods:** A hospital-based cross-sectional study was conducted, recruiting 317 participants for interviews using a proportional stratified sampling technique. The Composite International Diagnostic Interview (CIDI) was utilized to evaluate instances of suicide. The PHQ-9-(A) and an Oslo-3 social support instrument were used to assess depression and social support. Bivariate and multivariate binary logistic regressions were employed to identify factors associated with suicidal ideation and attempts. Statistical significance was indicated by a P-value <0.05, and the strength of association is presented as an adjusted odds ratio with a 95% confidence interval.

**Results:** The findings of this study revealed that the prevalence of suicidal ideation and suicide attempts were 12.6% and 6%, respectively. Among the respondents who had positive ideation, approximately 28% had thought about it at least once, 31.5% had thoughts at least two times, and 5% claimed they had contemplated committing suicide up to ten times in their lifetime. According to the multivariate analysis, being aged 15-19 years, having a widowed family status, a family history of mental illness, poor social support, and depression according to the PHQ-9 were significantly associated with suicidal ideation. Additionally, participants who had experienced a loss of a family member and those with depression had a significant risk of suicide attempts.

**Conclusion:** The results suggest that the magnitude of suicidal ideation and attempts is high, indicating a critical need for early suicide-focused regular screening for those on follow-up for other medical illnesses and linkage with mental health service providers.

## Introduction

Suicide originates from the Latin term for “self-murder” and signifies a person’s desire to die. There is a spectrum between contemplating suicide and carrying it out, with some individuals planning for an extended period before taking action, while others act on impulses without prior thought(1). It is a preventable public health problem worldwide, especially in adolescents. Adolescent suicidal behavior, which is an important public health problem worldwide, is a neglected public health issue, especially in middle and low-income countries Suicide is now the 10th leading cause of death worldwide and the 3rd leading cause of death between the ages of 15–24 years(2, 3).

Early adolescence is a period of significant change, during which children undergo physical changes associated with puberty and face new challenges in their transition to middle school where they encounter increased peer and academic pressures. Hence, thoughts about death and suicide become more common as children move through early adolescence (1). The age at which suicidal tendencies typically start is between 10 and 15 years old(4, 5).

Elements linked to a medical condition that can lead to both suicide and suicide attempts include reduced mobility, particularly when physical activity is crucial for work or leisure; physical disfigurement, especially among women; and persistent, unmanageable pain. Patients on hemodialysis are at high risk. In some cases, medications may cause depression and increase the risk of suicide. Among these drugs are corticosteroids, antihypertensive agents, and some anticancer agents. Alcohol-related illnesses, such as cirrhosis, are associated with increased suicide rates(1,5).

In the public, the annual global suicide rate is 11.4 per 100, 000 people, or 1 death every 40 seconds. In the United States, approximately 35,000 deaths are caused by suicide each year, which is approximately 100 deaths per day. It is estimated that for every completed suicide, there are 25 suicide attempts. While there have been notable changes in suicide rates among certain groups over the past century, the overall rate has remained relatively constant at approximately 12 per 100,000 people throughout the 20th and early 21st centuries. Internationally, suicide rates vary, with countries such as Lithuania, South Korea, Sri Lanka, Russia, Belarus, and Guyana having rates higher than 25 per 100,000 people, while countries such as Portugal, the Netherlands, Australia, Spain, South Africa, Italy, and Egypt have rates lower than 10 per 100,000 people(1).

Most suicides worldwide occur in low- and middle-income countries, where the majority of the global population resides. Regarding age, more than half of the global suicides occurred before the age of 45 years. Most adolescents who died by suicide 90% were from low and middle-income countries where nearly 90% of the world’s adolescents live(2). The prevalence of suicidal thoughts with a plan between both genders varied from 1.7% in the United Republic of Tanzania to 15.3% in Benin and Kenya (3). The African region had the highest overall pooled prevalence of suicidal ideation 21.6% and there was no evidence of sex differences. In contrast, suicide ideation in the region of the Americas was markedly greater in females than in males, with an estimated prevalence ratio of 1.7(2, 6).

In a meta-analysis and systematic review conducted in Ethiopia, it was found that the occurrence of suicidal thoughts and attempts varied between 1% to 55% and from 0.6% to 14%, respectively (7). Another study conducted among students in Ethiopia showed that the prevalence of suicidal thoughts and attempts ranged from 14% to 58.3% and from 4.4% to 7.4%, respectively (8).

In various studies conducted in Ethiopia from 2015 to 2022, significant rates of suicidal thoughts and attempts among adolescents and young adults were observed. In Dangila town (2015), 22.5% reported suicidal thoughts and 16.2% attempted suicide, with factors like school absenteeism and poor social support linked to these behaviors, while gender and alcohol use showed no correlation (9). In 2018, HIV-positive youth at SPHMMC and St. Peter Hospital in Addis Abeba reported 27.1% experiencing suicidal thoughts, with female gender, family death, advanced HIV stages, depression, and stigma being significant risk factors (12). A 2021 study in northwestern Ethiopia found that among substance users, 14.5% had suicidal thoughts and 9.9% attempted suicide, indicating a progression from thoughts to attempts (13). In 2022 a study in the Anywaa zone, in Gambela region of western Ethiopia, revealed that 62.3% of participants heard discussions about suicide, with many expressing feelings of hopelessness and belief that their families would be better off without them, highlighting a concerning trend in suicidal ideation and behaviors among young people in the region(11).

Although not all-suicidal ideation materializes into suicide attempts or suicide, it is the first step in the path to suicide. Therefore, this study was carried out to determine the prevalence and associated factors of suicide ideation and suicide attempt among adolescents especially in health care settings. Determining such information is useful for understanding the burden of mental health on adolescent health. This information may also be useful in targeting scarce mental health resources in the provision of interventions that reduce the occurrence of suicide in Ethiopia.

## Methodology

### Study setting

Addis Ababa is the capital city of Ethiopia. SPHMMC is a specialized hospital that is found in Addis Ababa. It is the leading teaching and tertiary referral hospital under the federal ministry of Ethiopia. Receiving around 200,000 patients each year and serving a catchment area of over 5 million people. The Pediatrics department services include inpatient, outpatient, emergency, intensive care and follow-up clinics. The regular Outpatient department provides services to more than 24,000 patients per year. In the pediatric department, there are approximately eleven specialty and subspecialty follow-up clinics led by subspecialists and general pediatricians with the addition of resident physicians, interns, nurses, runners, and data clerks. The study population included adolescents aged 10-19 years who visited the SPHMMC, Pediatrics Department, at the given three-month period to the Regular Outpatient Department and specialty follow-up clinics.

### Study period

August 1, 2023,G.C. to October 30, 2023, G.C.

### Study design

An Institution based cross-sectional study, with prospective data collection

### Study population

Adolescents aged 10-19 years who were receiving care at St. Paul’s Pediatrics Outpatient Department and Follow-up clinics, meeting the inclusion criteria.

### Inclusion criteria

Adolescents aged 10-19 years attending regular and follow-up clinics in the specified timeframe

### Exclusion criteria

1. Patients who were seriously ill or unable to communicate were excluded.
2. Patients who were receiving care primarily for suicide-related issues and
3. Patients who were already on established follow-up at Pediatric psychiatry clinic were excluded.

### Sample size

The single population proportion formula was used with a 5% margin of error, 95% confidence level, and 5% non-response rate; 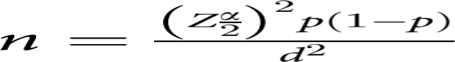

P= 27.1% -is the prevalence of suicidal ideation, which was taken from a similar study performed in a similar population background (12).

Z=1.96 at the α level, which is the standard score corresponding to the 95% confidence interval, and the d= margin of error is 0.05

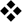 Therefore, based on the previous formula, N = (1.96)2 (0.271) (1-0.271)/ 0.0025 +15 = **317**

### Sampling technique

Participants were selected for interviews using the stratified sampling method, with their monthly average visits considered.

1. During the six months leading up to the study period, 2,892 patients visited the Pediatrics Outpatient Department, with 239 of them being adolescents aged 10-19 years. This indicates that adolescents represented approximately 8.2% of all cases during that period.
2. At the follow-up clinics, 9,540 patients were seen in the preceding six months. A two-level stratification was done. The first level determined the number of patients seen at each clinic, while the second level identified the number of adolescents treated at each clinic. The average percentage of patients from each clinic was calculated to project the number of patients to be screened for the research (Table 1).

**Table 1:**
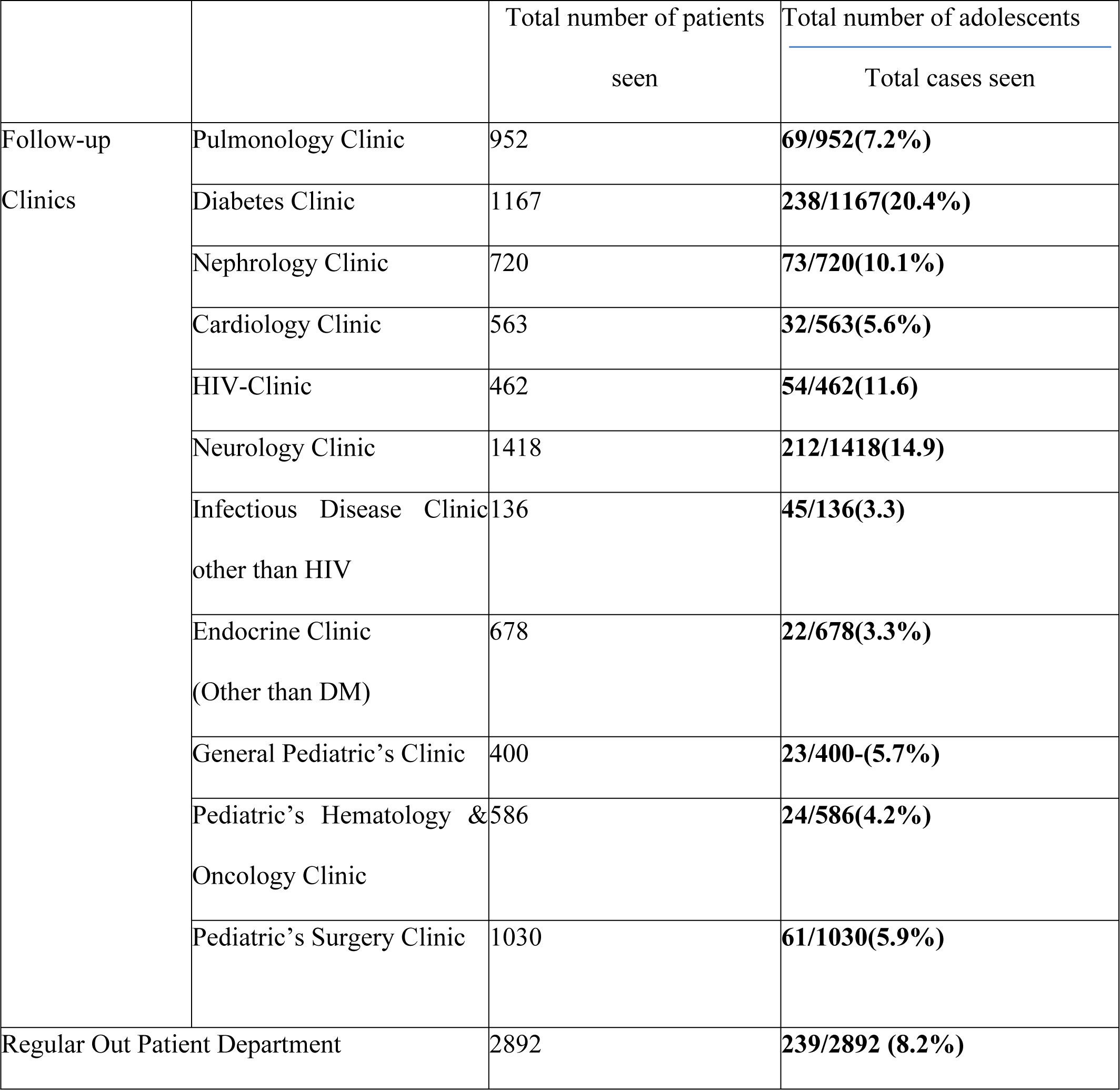
Percentage of study participants assigned based on the proportional stratified sampling method from each specialty follow-up clinic. * The overall total for the follow-up clinics has decreased from the previously mentioned figure because other specialty clinics in the hospital, such as the High-Risk Infant Clinic, do not include adolescents.

## Measurements

### Study variables

#### Independent variables

Age, Sex, Living arrangement, Homelessness, Family history of mental illness, Educational status, Parental Marital status, Family death, Presence of Social support, Depression, Parental educational status

#### Dependent variable

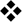 Suicidal ideation & Suicide attempt

## Instruments

Suicidal ideation and suicide attempts were assessed using the suicidality module of the World Mental Health (WMH) survey initiative version 3.0 of the World Health Organization (WHO) Composite International Diagnostic Interview (CIDI), which has been validated in Ethiopia(13,14). To evaluate suicidal thoughts, participants were asked the question: Have you ever thought about committing suicide? If “Yes”, the patient has suicidal ideation. For suicide attempts, participants were asked: ‘Have you ever tried to take your own life? If “Yes”, the patient has made a suicide attempt. The method of suicide attempt and reason for the attempt was assessed. Depression was measured by a scoring system that is modified for adolescents PHQ-9-(A) (14). A cutoff score of ten or higher was utilized to indicate depression. Social support was measured by the Oslo-3 social support scale(15). The scale ranged from 3–14, and scores of 3–8, 9–11, and 12–14 indicated “poor”, “moderate” and “strong” social support, respectively. To assess current substance use, respondents were asked: Have you ever used a substance (nonmedical use) in your lifetime? In addition, if the response was yes, further questions were asked on the specific substance and frequency of its use. Items on sociodemographic factors (age, sex, ethnicity, religion, marital status, educational status, and occupational status) were adopted from a variety of studies.

## Data collection

The data was collected through face-to-face interviews via structured questionnaire by trained physicians using the local language (Amharic) version of the tool. The data was collected by physicians trained on the introduction to suicide, and associated factors and skills related to how to assess this sensitive issue, interviewing skills, sampling, recruitment, and ethical aspects of the research. The data was collected prospectively and participants who displayed positive suicide ideation and attempt were linked to pediatrics psychiatry clinic for further evaluation and management.

## Data Processing and Analysis

All the data were checked for completeness and consistency and entered into Google forms, which were subsequently coded and exported to SPSS version 26.0 for analysis. A descriptive and bivariate logistic regression analysis was used to compute the frequency distribution and to test the association between independent and dependent variables, respectively. Factors associated with suicidal ideation and suicide attempts were selected during the bivariate analysis with a p-value <0.05 for further multivariate analysis, in which variables with a P-value less than 0.05 at the 95% confidence interval were considered statistically significant.

## Operational definition

**Suicide**: - Self-inflicted death with explicit or implicit evidence that the person intended to die.

**Suicidal ideation**: - Thought of serving as the agent of one’s death; seriousness may vary depending on the specificity of the suicidal plan and the degree of suicidal intent.

**Suicidal intent**:-Subjective expectation and desire for a self-destructive act to end in death.

**Suicidal attempt**: - self-injurious behavior with a nonfatal outcome accompanied by explicit or implicit evidence that the person intended to die.

**Adolescents**: - defined as individuals between 10 and 19 years of age (17).

## Data Quality Control and Management

To ensure the quality of the data, structured data was collected. The investigator trained the data collector. Each day, the data was checked for accuracy and completeness. The participants’ names were not collected and the data were de-identified after the questionnaire was completed. The data were then stored in a secured location and only the investigator had access to this location.

## Ethical clearance

Ethical approval was obtained from the SPHMMC-IRB (institutional review board), and confidentiality of the information was ensured using codes rather than names during the registration of the data. Informed and written consent was obtained from the parents or caretakers, and assent was obtained from the adolescents. Participants who displayed depression, positive suicidal ideation and attempted suicide were linked to a pediatrics psychiatry clinic to receive appropriate medical and psychological care.

## Results

### Sociodemographic characteristics of the study participants (n=302)

A total of 302 of the estimated 317 adolescents participated in the study, resulting in a 95.1% response rate. Fifteen participants, or 4.9%, did not respond to all of the questions and were therefore excluded from the analysis. Fifty-three percent of the study participants were in the age group of 10-14 years and 53% were female. 71.9% of the participants were from Addis Ababa, and 79.8% of the parents of the participants were married. Seventy-nine percent of the participants had a secondary education, and 47.7% of the families had completed secondary education. Nearly 23% of the participants reported the loss of at least one family member, while 10.6% of the participants had a family member with a mental illness. The vast majority of individuals in the study, 99%, were residing with their families, whereas only small percentages, 1%, of the children were without a home (Table 2).

**Table 2.** Sociodemographic characteristics of the adolescents attending SPHMMC, Addis Abeba, Ethiopia, 2023 (n=302)

| Variable | Frequency | Percent |
| --- | --- | --- |
| **Age in years** |  |  |
| 10-14 | 160 | 53 |
| 15-19 | 142 | 47 |
| **Sex** |  |  |
| Female | 160 | 53.0 |
| male | 142 | 47.0 |
| **Religion** |  |  |
| Muslim | 86 | 28.5 |
| Orthodox Christian | 153 | 50.7 |
| Protestant Christian | 63 | 20.9 |
| **Residence** |  |  |
| Addis Ababa | 217 | 71.9 |
| Outside of Addis Abeba | 85 | 28.1 |
| **Parental Marital status** |  |  |
| Married | 243 | 80.4 |
| Divorced | 41 | 13.6 |
| Widowed | 18 | 6.0 |
| **Educational status** |  |  |
| Illiterate | 1 | 0.3 |
| Primary | 63 | 20.9 |
| Secondary | 237 | 78.5 |
| College and above | 1 | 0.3 |
| **Parental Educational status** |  |  |
| Illiterate | 37 | 12.3 |
| Primary | 54 | 17.9 |
| Secondary | 144 | 47.7 |
| College and above | 67 | 22.2 |
| **Family death** |  |  |
| Yes | 69 | 22.8 |
| No | 233 | 77.2 |
| **Family history of mental illness** |  |  |
| Yes | 32 | 10.6 |
| No | 270 | 89.4 |
| **Family history of mental illness** |  |  |
| Yes | 32 | 10.6 |
| No | 270 | 89.4 |
| **Living arrangement** |  |  |
| Alone | 3 | 1.0 |
| With family | 299 | 99.0 |
| **Homelessness** |  |  |
| Yes | 3 | 1.0 |
| No | 299 | 99.0 |

### Characteristics of the Oslo-3 social support scale (OSSS)

Forty five percent of the individuals in the study had a support system of 3-5 people whom they could rely on during times of significant difficulty, 54% of the participants disclosed that people show some degree of concern and interest towards them, and approximately 22.8% of the participants reported some degree of difficulty getting help from their neighbors(Fig 1).

**Figure 1.**
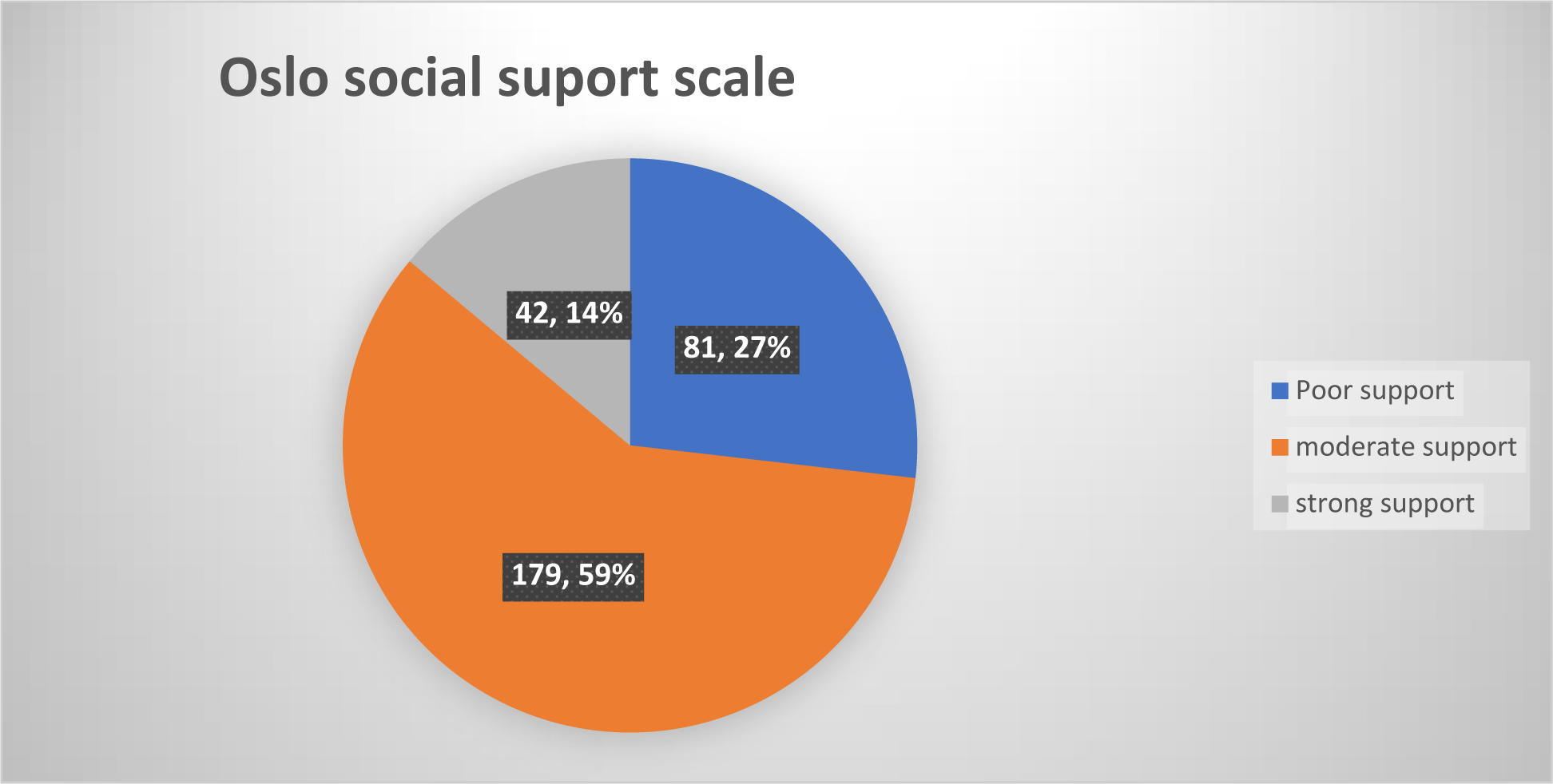
Social support of the adolescents attending SPHMMC, Addis Abeba, Ethiopia, 2023 (n=302)

### Depression related characteristics of the study participants

Twelve of the participants (4%) had a positive screen for depression according to the PHQ-9 (A) assessment. Regarding the specific assessment variable, 81.1% of the participants did not experience symptoms of feeling down, depressed, irritable, or hopeless. 83.8% of the participants did not report any loss of interest, and 91.1% of the participants did not experience any trouble concentrating on things such as schoolwork, reading, or watching TV. Eighteen percent of the study participants felt depressed or sad on most days, but approximately 87.7% of the participants did not complain of any difficulty doing their work or taking care of things at home.

### Substance abuse related characteristics

Approximately 5% of the study participants reported ever using recreational drugs during their lifetime. Of the participants, 78.6% reported using alcohol at least once, while 14.3% reported using Khat (Table 3).

**Table 3.** Substance abuse related use among the adolescents attending SPHMMC, Addis Abeba, Ethiopia, 2023 (n=302)

| Have you ever used recreational drugs in your lifetime? |  |  |
| --- | --- | --- |
| Yes | 14 | 4.6 |
| No | 288 | 95.4 |
| **If yes types of drugs (n=14)** |  |  |
| Alcohol drinking | 11 | 78.6 |
| Khat Chewing | 2 | 14.3 |
| Glue sniffing (Mastish) | 1 | 7.1 |

‘‘Khat’’- is a stimulant plant, Mastish-local term for ‘‘glue sniffing’’

### Prevalence of suicidal ideation and attempt

The prevalence rates of suicidal ideation and suicide attempts were 12.6% (25 female, 13 males) and 6% (10 females, 8 males) respectively (Fig. 2). Among the respondents who had positive ideation approximately 28% had thought about it at least once, 31.5% of them had thoughts at least 2 times, and 5% of the study participants claimed they had thought about committing suicide up to 10 times in their lifetime. Six percent of the study participants had attempted suicide, of whom eight (44.4%) attempted using drugs, four (22.4%) attempted using poisons other drugs, and 16.7% used a knife/sharp objects or hanging as a means of ending their life. Of those 18 participants who attempted suicide, five said that the death of a family member was their reason for the attempt, four disclosed family conflict, two said they felt they did not have any way out, and the other two said because of academic failure.

**Figure 2.**
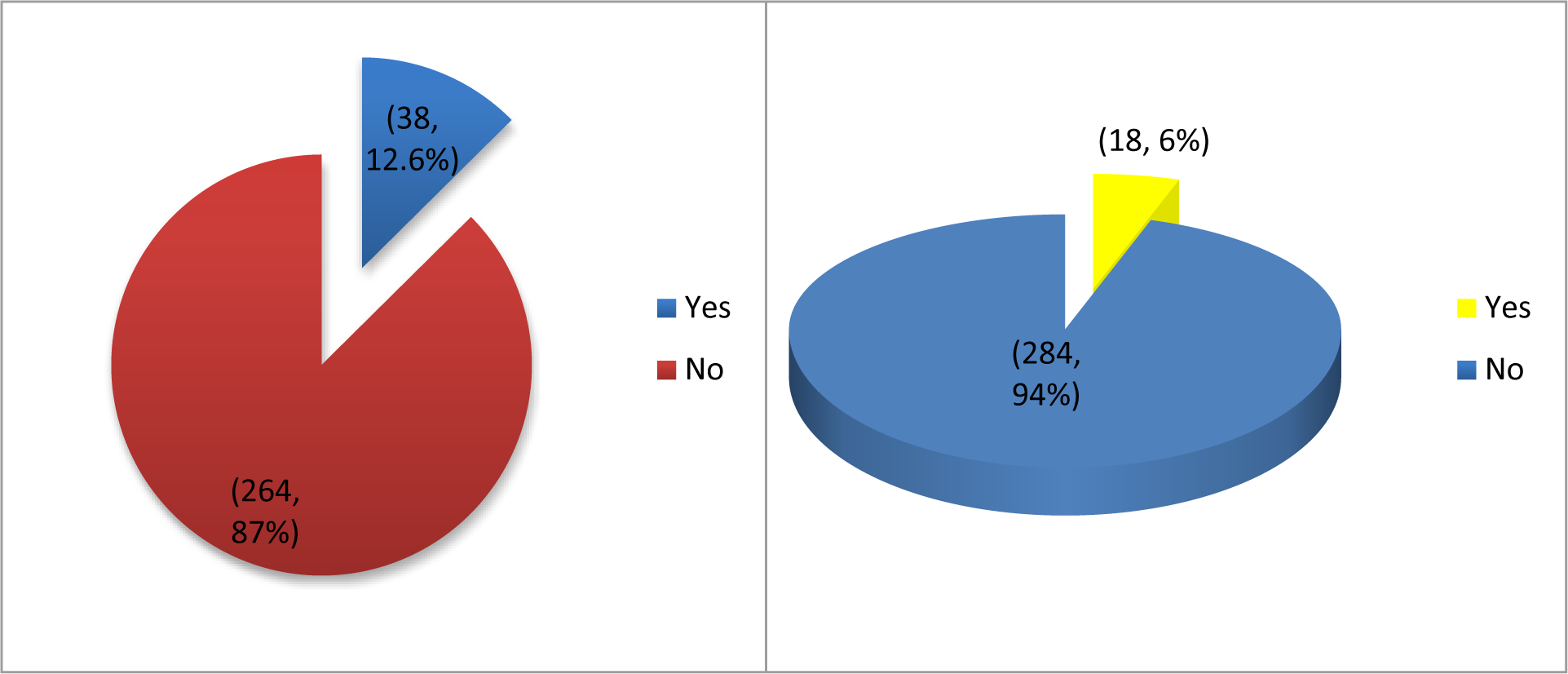
Prevalence of suicidal ideation (First pie chart) & Prevalence of suicide attempt (Second pie chart) of the adolescents attending SPHMMC, Addis Abeba, Ethiopia, 2023 (n=302)

In this study, despite the fact both suicide ideation and suicide attempt were numerically greater among girls than among boys, but on multivariate logistic regression gender didn’t show statistically significant association with suicidal ideation and attempt (P-0.063).

Concerning the seriousness of suicide attempts, three (17%) of the study participants reported having attempted suicide to “cry for help”, while nine (50%) reported a serious attempt to kill themselves, and it was only luck that they have not succeeded (Table 4).

**Table 4.** Prevalence of suicidal ideation and attempt among the adolescents attending SPHMMC, Addis Abeba, Ethiopia, 2023 (n=302).

| Variable | Frequency | Percent |
| --- | --- | --- |
| **Have you ever thought about committing suicide?** |  |  |
| No | 264 | 87.4 |
| Yes | 38 | 12.6 |
| **Number of thoughts committing suicide (n=38)** |  |  |
| 1 | 9 | 23.7 |
| 2 | 12 | 31.6 |
| 3 | 9 | 23.7 |
| 4 | 2 | 5.3 |
| 5 | 1 | 2.6 |
| 6 | 2 | 5.3 |
| 8 | 1 | 2.6 |
| 10 | 2 | 5.3 |
| **Have ever thought about committing suicide in the past month?** |  |  |
| Yes | 10 | 3.3 |
| No | 292 | 96.7 |
| **Have you ever attempted suicide** |  |  |
| Yes | 18 | 6.0 |
| No | 284 | 94.0 |
| **Mechanism of attempted suicide (n=18)** |  |  |
| Drugs | 8 | 44.4 |
| Hanging | 3 | 16.7 |
| Knives or other sharp objects | 3 | 16.7 |
| Poisoning other than drugs | 4 | 22.2 |
| **Have you ever attempted suicide in the past month** |  |  |
| Yes | 17 | 5.6 |
| No | 285 | 94.4 |
| **Reason for attempt (n=17)** |  |  |
| Academic failure | 2 | 11.8 |
| Death of a family member | 5 | 27.8 |
| Family conflict | 4 | 23.5 |
| Feel like there is no other way out | 2 | 11.8 |
| Insult by her friend in school | 1 | 5.9 |
| Left from previous school and friends, feels the previous school understood her more, and are abusive at the current school | 1 | 5.9 |
| Medical Illness | 1 | 5.9 |
| School conflict with a friend | 1 | 5.9 |
| **Are you currently experiencing thoughts of self-harm?** |  |  |
| Yes | 6 | 2.0 |
| No | 296 | 98.0 |

### Determinant factors for suicidal ideation

Accordingly, bivariate logistic regression revealed that age, sex, parental marital status, parental education level, death of a family member, family history of mental illness, poor social support and depression were associated with suicidal thoughts. Multivariate logistic regression revealed that study participants aged 15-19 years had a 5.2-fold greater likelihood of having suicidal ideation than did those aged 10-14 years (AOR=5.2, 95% CI=1.44, 18.73), and study participants whose families were widowed had a 6.6-fold greater likelihood of having suicidal ideation than did those whose families were married (AOR=6.6, 95% CI=1.08, 40.63). The study participant who had experienced death of a family member had a 4.2 folds greater likelihood of having suicidal thoughts compared to those who did not (AOR=4.2, 95%CI=1.04, 17.08) and the study participant who had a family history of mental illness had 7.6 folds increase in having suicidal thoughts compared to their counterparts (AOR=7.6, 95%CI=1.82, 32.05).

Compared with participants who had strong support, participants who had poor social support had a 2.9 fold increase in suicidal ideation (AOR=2.9, 95%CI=1.60, 14.70), and participants who had depression according to the PHQ9(A) had a 14.1 fold increase in suicidal ideation compared to those who did not (AOR=14.1, 95%CI=1.82, 39.31), additionally, participants who had depression or felt sad on most of the days in the past year had a 13.8 fold increase in suicidal ideation compared to those with no such symptoms (AOR=13.8, 95%CI=1.82, 47.93)(Table 6).

**Table 5:** Cross-table relationship between the site of evaluation and whether the participants had attempted suicide.

| Site of Evaluation | Have you ever attempted suicide |  |  |
| --- | --- | --- | --- |
|  | Yes | No | Total |
| Cardiac Clinic | 0 | 16 | 16 |
| Pulmonology Clinic | 0 | 22 | 22 |
| Diabetes Clinic | 0 | 62 | 62 |
| Endocrine other than Diabetes | 0 | 5 | 5 |
| General Pediatrics Clinic | 2 | 16 | 18 |
| Hematology & Oncology Clinic | 1 | 12 | 13 |
| Infectious Disease Clinic(Other than HIV) | 0 | 9 | 9 |
| HIV-Clinic | 11 | 24 | 35 |
| Neurology Clinic | 1 | 44 | 45 |
| Pedi-Surgery Clinic | 0 | 21 | 21 |
| Nephrology Clinic | 0 | 31 | 31 |
| Regular Outpatient department | 2 | 22 | 25 |
| **Total** | **18** | **284** | **302** |

**Table 6.** Factors associated with a suicide ideation of participants at SPHMMC, Addis Abeba, Ethiopia, 2023 n=302.

| Variable | Thought committing suicide |  | P-value | COR with 95%CI | P-value | AOR with 95%CI |
| --- | --- | --- | --- | --- | --- | --- |
|  | Yes | No |  |  |  |  |
| Age in years |  |  |  |  |  |  |
| 10-14 | 9 | 133 | 1 |  |  |  |
| 15-19 | 29 | 131 | 0.003 | 3.3(1.49, 7.18) | 0.012 | **5.2(1.44, 18.73)** |
| **Sex** |  |  |  |  |  |  |
| Female | 25 | 135 | 0.094 | 1.8(0.90, 3.75) | 0.063 | 3.1(0.94, 10.51) |
| Male | 13 | 129 | 1 |  |  |  |
| **Parent marital status** |  |  |  |  |  |  |
| Married | 15 | 219 | 1 |  |  |  |
| Divorced | 9 | 32 | 0.002 | 4.1(1.66, 10.16) | 0.181 | 2.7(0.63, 12.09) |
| Single | 4 | 5 | 0.001 | 11.7(2.84, 48.08) | 0.101 | 6.4(0.66, 62.89) |
| Widowed | 10 | 8 | 0.000 | 18.3(6.28, 53.03) | 0.041 | **6.6(1.08, 40.63)** |
| **Parent education level** |  |  |  |  |  |  |
| Illiterate | 10 | 27 | 0.019 | 3.8(1.24, 11.41) | 0.971 | 0.96(0.13, 7.25) |
| Primary | 7 | 47 | 0.481 | 1.5(0.48, 4.81) | 0.503 | 0.52(0.08, 3.55) |
| Secondary | 15 | 129 | 0.742 | 1.2(0.44, 3.19) | 0.952 | 1.1(0.21, 5.23) |
| College and above | 6 | 61 | 1 |  |  |  |
| **Family died** |  |  |  |  |  |  |
| Yes | 22 | 47 | 0.000 | 6.3(3.09, 13.00) | 0.044 | **4.2(1.04, 17.08)** |
| No | 6 | 217 | 1 |  | 1 |  |
| **Family history of mental illness** |  |  |  |  |  |  |
| Yes | 14 | 18 | 0.000 | 7.9(3.53, 18.01) | 0.005 | **7.6(1.81, 32.05)** |
| No | 24 | 246 | 1 |  | 1 |  |
| **OSLO social support scale** |  |  |  |  |  |  |
| Poor | 22 | 59 | 0.030 | 3.5(1.13, 11.08) | 0.018 | **2.9(1.60, 14.70)** |
| Moderate | 12 | 167 | 0.528 | 0.68(0.21, 2.23) | 0.949 | 1.1(0.19, 5.69) |
| Strong | 4 | 38 | 1 |  | 1 |  |
| **Depression using PHQ9** |  |  |  |  |  |  |
| Yes | 10 | 2 | 0.000 | 16.8(9.76, 24.29) | 0.011 | **14.1(1.82, 39.31)** |
| No | 28 | 262 | 1 |  | 1 |  |
| **In the past year felt depressed or sad most of the days** |  |  |  |  |  |  |
| Yes | 29 | 25 | 0.000 | 13.8(3.12, 72.35) | 0.000 | **13.8(3.97, 47.93)** |
| No | 9 | 239 | 1 |  | 1 |  |

### Determinant factors for suicide attempts

In this study, family death, family history of mental illness, poor social support and the presence of depression were associated with suicide attempt risk according to bivariate logistic regression. Multivariate logistic regression revealed that suicide attempts were 10.4 fold increase in participants who have experienced loss of a family member than those that did not (AOR=10.4, 95%CI=3.10, 36.49), and study participants who had depression according to the PHQ9 (A) had a 12.6 fold increase in attempting suicide compared to those that did not (AOR=12.6, 95%CI=2.42, 65.77)(Table 7).

**Table 7.** Factors associated with a suicide attempt of the adolescents attending SPHMMC, Addis Abeba, Ethiopia, 2023 (n=302).

| Variable | Have you ever attempted suicide |  | P-value | COR with 95%CI | P-value | AOR with 95%CI |
| --- | --- | --- | --- | --- | --- | --- |
|  | Yes | no |  |  |  |  |
| Age in years |  |  |  |  |  |  |
| 10-14 | 6 | 136 | 1 |  | 1 |  |
| 15-19 | 12 | 148 | 0.236 | 1.8(0.67, 5.03) | 0.468 | 0.64(0.19, 2.15) |
| **Family death** |  |  |  |  |  |  |
| Yes | 12 | 56 | 0.000 | 10.6(3.62, 30.92) | 0.000 | **10.4(3.10, 36.49)** |
| no | 5 | 228 | 1 |  | 1 |  |
| **Family history of mental illness** |  |  |  |  |  |  |
| Yes | 6 | 26 | 0.003 | 4.9(1.72, 14.32) | 0.233 | 2.3(0.59, 8.77) |
| no | 12 | 258 | 1 |  | 1 |  |
| **OSLO social support scale** |  |  |  |  |  |  |
| Poor | 12 | 69 | 1 |  | 1 |  |
| Moderate | 5 | 174 | 0.001 | 0.17(0.06, 0.49) | 0.097 | 6.2(0.72, 54.02) |
| strong | 1 | 40 | 0.064 | 0.14(0.02, 1.12) | 0.612 | 1.8(0.19, 16.67) |
| **Presence of depression** |  |  |  |  |  |  |
| Yes | 5 | 7 | 0.000 | 15.2(1.25, 54.48) | 0.003 | **12.6(2.42, 65.77)** |
| No | 13 | 277 | 1 |  | 1 |  |

## Discussion

The findings of this study revealed that the prevalence of thought about committing suicide and attempting suicide were 12.6 % and 6 % respectively. This finding was in line with a study done in the Republic of Tanzania, Benin and Kenya, and with the Ethiopian meta-analysis (2, 6, 7). This study is also in line with the study performed in northwestern Ethiopia in Gonder (13). This finding was lower than that of another study performed at SPHMMC and St. Peters hospital (12) and African pooled prevalence (6). This variation could be explained by the smaller sample size used in the study and differences in study settings, since most of the studies were conducted in a school setting rather than in a health care setting.

Men commit suicide more than four times as often as women, regardless of age or race, in the United States even though women attempt suicide or have suicidal thoughts three times as often as men do(1). In this study, despite the fact both suicide ideation and suicide attempt were numerically greater among girls than among boys, multivariate logistic regression gender didn’t show statistically significant association with suicidal ideation and attempt (P-0.063). This finding is in line with studies conducted in sub-Saharan countries (3), other Ethiopian studies such as those in the town of Dangla (9), and studies conducted among university students in Ethiopia (8) which showed gender has no significance with ideation or attempt of suicide.

In this study, participants aged 15-19 years had 5.2 fold greater suicidal ideation than did those aged 10-14 years (AOR=5.2, 95%CI=1.44, 18.73). This might be because Suicide attempts are uncommon in children under 12 because their cognitive development provides a protective barrier (1); additionally adolescents and young adults have a more advanced level of cognitive development than younger children do. They may have a better understanding of death and its permanence, as well as the ability to plan and execute suicide attempts (1, 4).

The study participants from widowed families had a 6.6 fold greater incidence of suicidal ideation than did those from families with both parents (AOR=6.6, 95%CI=108, 40.63). This might be because the loss of a parent can result in financial instability for the family, especially if the deceased parent was the primary breadwinner. Financial stressors such as difficulty paying bills, affording necessities, or experiencing housing instability can increase children’s anxiety and depression, elevating their risk of suicide attempts.

The study participants with poor social support had a 2.9 fold greater risk of having suicidal ideation than did those with strong social support (AOR=2.9, 95%CI=1.60, 14.70). This finding is in line with other studies conducted in Ethiopia, such as the Fiche town study (10), Dangla Town study (9), Gonder University study (8), LMIC study (3,6); the St.Pauls and St. peter joint study (12). This could be explained by receiving social support enhances a sense of belongingness and ultimately decreases the likelihood of suicidal thoughts (8). One other potential explanation for this connection may be that experiencing neglect from neighbors, friends, family, and others can greatly contribute to feelings of hopelessness and low self-worth, ultimately leading to suicidal thoughts especially in young people (1,5).

Participants with depression according to the PHQ 9(A) had 14.1 fold greater risk of having suicidal thoughts than did those that did not (AOR=14.1, 95%CI=1.82, 39.31). This finding is also in line with most of the studies mentioned above (3, 6, 10-12). This may be because individuals with depression often have reduced levels of serotonin, a neurotransmitter. This imbalance in the brain could lead to feelings of hopelessness, guilt, and worthlessness, which may increase the risk of suicidal thoughts (5,16,18). Additionally, respondents with a family history of mental illness had over seven times the likelihood of experiencing suicidal thoughts. This observation aligns with another study that found a correlation between a family history of psychiatric disorders and an increased risk of suicidal ideation (8, 19). However, it is crucial to highlight that the assessment of family mental health history in this study was based on a single question instead of utilizing formal diagnostic tools.

Suicidal ideation, suicide attempts, and completed suicides in children are serious concerns that require immediate attention and intervention. It is essential for parents, caregivers, educators, and healthcare providers to be aware of warning signs that may indicate a child is experiencing suicidal thoughts.

## Limitations

Our cross-sectional design prevented us from reporting causal relationships of the associations we found. In addition, social desirability and recall bias might have also been the other limitation. Since the data was collected through face-to-face interviews, participants may have provided socially acceptable responses, particularly regarding questions about substances. The other limitation of the study is that it is a single institution study; the results cannot be generalized to the general population.

## Conclusion

The magnitude of suicidal ideation and attempts among adolescents was found to be high. Suicidal ideation was statistically significant in those aged 15-19 years, those with Poor social support, those with a family history of mental illness; those who lost a close family member and those with depression. However, suicidal attempts were more common among those who had experienced family death and those with depression.

## Recommendation

We suggest implementing regular screening for suicide risk at an early stage and connecting individuals with mental health service providers. Consider screening parameter for adolescents during follow up visit for depression, substance use and suicidal ideations.

Adolescents with chronic medical illnesses are at greater risk for suicidal thoughts and behaviors. To reduce this risk, we recommend providing comprehensive mental health support, educating healthcare providers, foster a multidisciplinary approach, support peer connections, monitoring medication side effects, involving parents and caregivers, and providing ongoing support. The Last recommendation is regarding follow-up, strengthening ongoing support and follow-up for adolescents who have previously shown signs of suicidal ideation or attempted suicide. This can include regular check-ins, counseling sessions, and the involvement of mental health professionals.

## Data Availability

All relevant data are within the paper and its Supporting Information files.

## DECLARATION

### Ethical considerations

Ethical approval was obtained from the SPHMMC-IRB (institutional review board), and confidentiality of the information was ensured using codes rather than names during the registration of the data. Informed and written consent was obtained from the parents or caretakers, and assent was obtained from the adolescents. Participants who displayed depression, positive suicidal ideation and attempted suicide were linked to pediatrics psychiatry clinic to receive appropriate medical and psychological care.

### Competing interests

The authors declare that they have no competing interests.

### Funding

Funding was from the SPHMMC.

### Authors’ Contributions

Conceptualization: Tigist Bacha

Data Curation: Dawit Sintayehu, Tigist Bacha

Formal analysis: Dawit Sintayehu

Funding acquisition: Dawit Sintayehu

Investigation: Dawit Sintayehu, Tigist Bacha

Methodology: Dawit Sintayehu, Tigist Bacha, Tigist Zerihun

Supervision: Tigist Bacha, Tigist Zerihun

Writing-original draft: Dawit Sintayehu

Writing-review & editing: Dawit Sintayehu, Tigist Bacha, Tigist Zerihun

### Availability of data and materials

All data generated or analyzed during this study are included in this manuscript and the supplementary section.

## Acknowledgements

The authors would like to acknowledge St. Paul’s Hospital Millennium Medical College (SPHMMC), Department of Pediatrics and Child Health and the study participants for their dedicated cooperation and our data collectors for their valuable time and effort. We would like to thank the SPHMMC IRB for providing ethical clearance and Dr. Yossef Alnasser for his valuable and critical feedback.

## Acronyms

ART: Antiretroviral therapy
AOR: Adjusted Odds Ratio
CDC: Center for disease control
CI: Confidence interval
CIDI: Composite International Diagnostic Interview
DM: Diabetes mellitus
DSM: Diagnostic and Statistical Manual of Mental Disorders
HIV: Human immunodeficiency virus
N: Total Sample size
OSSS: Oslo-3 Social Support Scale
PHQ-9-(A): Patient Health Questioner for adolescents
PHO: Pediatrics Hemato-oncology
PTSD: Post traumatic stress disorder
ROPD: Regular Out Patient Department
SPHMMC: St. Paul’s hospital millennium medical college
SPSS: Statistical package for social sciences
WHO: World Health Organization
WMH: World Mental Health

## Notes

### Competing Interest Statement

The authors have declared no competing interest.

### Author Declarations

This research has been approved by the St. Paul's Hospital Millennium Medical College IRB Reference no. pm13/916, on 13/6/2013. the IRB file is attached with the main manuscript during submission. We have prospectively recruited human participants and collected data for the study using face to face interview using structured questionnaires. Participants provided informed and verbal assent. It was witnessed and documented in the data forms. We have obtained consent from parents or guardians.

